# Let the DOCTOR Decide Whom to Test: Adaptive Testing Strategies to Tackle the COVID-19 Pandemic

**DOI:** 10.1101/2020.12.18.20248498

**Authors:** Yu Liang, Amulya Yadav

## Abstract

A robust testing program is necessary for containing the spread of COVID-19 infections before a vaccine becomes available. However, due to an acute shortage of testing kits (especially in low-resource developing countries), designing an optimal testing program/strategy is a challenging problem to solve. Prior literature on testing strategies suffers from two major limitations: (i) it does not account for the trade-off between testing of symptomatic and asymptomatic individuals, and (ii) it primarily focuses on static testing strategies, which leads to significant shortcomings in the testing program’s effectiveness. In this paper, we address these limitations by making five novel contributions. (i) We formally define the optimal testing problem and propose the DOCTOR POMDP model to tackle it. (ii) We solve the DOCTOR POMDP using a scalable Monte Carlo tree search based algorithm. (iii) We provide a rigorous experimental analysis of DOCTOR’s testing strategies against static baselines - our results show that when applied to the city of Santiago in Panama, DOCTOR’s strategies result in ∼40% fewer COVID-19 infections (over one month) as compared to state-of-the-art static baselines. (iv) In addition, we analyze DOCTOR’s testing policy to derive insights about the reasons behind the optimality of DOCTOR’s testing policy. (v) Finally, we characterize conditions (of the real world) under which DOCTOR’s optimization would be of most benefit to government policy makers, and thus requires significant attention from researchers in this area. Our work complements the growing body of research on COVID-19, and serves as a proof-of-concept that illustrates the benefit of having an AI-driven adaptive testing strategy for COVID-19.

## 1 INTRODUCTION

COVID-19 (or coronavirus) is an urgent public health crisis - within nine months, COVID-19 has infected more than 35 million people, and has resulted in ∼ 1 million deaths worldwide [42]. In fact, it has been declared as a global pandemic by the World Health Organization (WHO) [11]. Unfortunately, despite the enforcement of stringent preventive measures, the spread of COVID-19 still does not appear to be slowing down.

According to the Centers for Disease Control and Prevention (CDC), there is currently no vaccine to prevent COVID-19, and the best way to prevent infection is to avoid being exposed to the COVID-19 virus through social distancing [10]. In response to this outbreak, governments around the world have enacted several unprecedented “aggressive social distancing” measures, e.g., schools and businesses have been closed, and people have been ordered to stay home. Although these measures are crucial for slowing down infections and preventing healthcare systems from getting overwhelmed, it is difficult to enforce them for a prolonged period of time, due to the devastating impact of these orders on peoples’ livelihoods. This negative impact is most acutely felt in developing countries, where a majority of the population rely on daily wages for a living, and prolonged stay-at-home orders cut off the sole means of sustenance for this population. In fact, more than 71 million people have been pushed to extreme poverty due to social distancing measures enforced in developing countries [39]. Thus, governments (in developing countries) are under severe pressure to remove social distancing measures and quarantine protocols as soon as possible. However, the hasty removal of quarantine measures may only serve to exacerbate the COVID-19 public health crisis. Therefore, finding innovative science-backed methods to contain the spread of COVID-19 is now of the utmost importance.

A robust COVID-19 testing program is necessary for containing the spread of infections, as it can: (i) help identify and quarantine infected patients, which can break the chain of COVID-19 transmissions and reduce the total number of infections; and at a higher level, (ii) aggregate results from COVID-19 testing programs can help epidemiologists and policy makers in determining where communities/states/countries are on the epidemic curve, which enables them to take more well-informed decisions about the removal of stay-at-home orders.

However, designing the optimal testing program for COVID-19 is a challenging problem because of three major reasons. First, in addition to testing individuals with symptoms who show up at the hospital (i.e., symptomatic testing), the CDC also recommends testing individuals without symptoms in the public (i.e., asymptomatic testing) in order to detect COVID-19 early and stop transmission quickly [8]. Second, policy makers (especially in developing countries) are constrained in the number of tests (both symptomatic and asymptomatic) that they can conduct on a daily basis, and thus, they need to strategically allocate their limited number of tests among symptomatic and asymptomatic patients. Finally, an optimal testing strategy needs to be adaptive, as the number of symptomatic/asymptomatic tests per day should be increased/decreased adaptively depending on the number of positively diagnosed people in previous days of testing. Therefore, policy makers need to intelligently allocate their limited resources (i.e., limited number of COVID-19 testing kits) over a prolonged period of time in order to minimize the total (cumulative) number of COVID-19 infections.

To this date, while almost every country has a COVID-19 testing strategy in place, these strategies are mostly static (i.e., non-adaptive), potentially causing significant shortcomings in their effectiveness in containing COVID-19 (we validate this in our experimental analysis). In this paper, we overcome this limitation via three novel contributions. First, we provide a formal definition of the optimal testing problem and propose the DOCTOR (**D**esign of **O**ptimal **C**OVID-19 **T**esting **Or**acle) model, which casts the optimal testing problem as a Partially Observable Markov Decision Process (POMDP). Second, we solve DOCTOR’s POMDP model using a Monte Carlo tree search based algorithm [33]. Our POMDP based algorithm has three key novelties: (i) it models the spread of the COVID-19 virus via SEIR model dynamics [2]; (ii) it optimally trades off between the amount of resources (i.e., testing kits) that should be invested in symptomatic versus asymptomatic testing to find the optimal testing strategy; and (iii) our POMDP model adaptively updates its future long term policy based on aggregate testing results (i.e., how many symptomatic and asymptomatic tests came out positive, etc.) from previous rounds. Finally, and most importantly, we also provide a rigorous experimental analysis of DOCTOR’s testing strategy against static testing programs to illustrate the effectiveness of our approach. Our experiments reveal that DOCTOR’s testing strategy was able to outperform state-of-the-art baselines by achieving ∼ 40% fewer COVID-19 infections, when applied to city of Santiago, Panama. The result illustrates the benefit of having an adaptive strategy. In addition, we analyze DOCTOR’s testing policy to derive insights about the reasons behind the optimality of DOCTOR’s testing policy. At the end, we also provide a characterization of the problem conditions in which it is most advisable to rely on DOCTOR for determining the COVID-19 testing policy.

COVID-19 is the greatest public health crisis that the world has experienced in the last century. Tackling it requires the collective will of experts from a variety of disciplines. While a lot of efforts have been made by AI researchers in developing agent-based models for simulating the transmission of COVID-19 [21, 41, 44], we believe that AI’s enormous potential can (and should) be leveraged to design decision support systems (e.g., in the allocation of limited healthcare resources such as testing kits) which can assist epidemiologists and policy makers in their fight against this pandemic. Our work represents the first step in developing such a decision support system, and should serve as a proof-of-concept that illustrates the benefit of having an AI-driven adaptive testing strategy.

## 2 RELATED WORK

To the best of our knowledge, very little prior work on COVID-19 has focused on resource allocation problems that arise due to acute shortages of COVID-19 testing kits (despite this shortage being an unfortunate reality in many developing countries, such as Nepal [18]). Nevertheless, we discuss prior work in three related areas.

First, we describe prior work on optimizing COVID-19 testing strategies. A lot of work has focused on optimizing group (pooled) testing techniques, in which COVID-19 tests are conducted on blood samples pooled from several patients in order to reduce the number of tests required to diagnose a population [4, 28, 37]. This body of research is complementary to our work, as DOCTOR’s model employs group testing to conduct tests on asymptomatic individuals, so it can exploit performance gains due to these optimized group testing techniques. Also, Singh et al. [35] proposed POMDP based sequential testing strategies to optimize contact tracing, i.e., given a confirmed COVID-19 positive diagnosis, which of their contacts should be tested next. However, DOCTOR looks at a complementary goal - given a limited number of testing kits, how to optimally distribute them among symptomatic and asymptomatic individuals.

Second, we describe prior work on epidemiological models for simulating COVID-19 transmissions. A lot of efforts have been made by AI researchers to develop SEIR type models and agent based models which can simulate the progression of COVID-19 in a population of susceptible individuals [12, 31, 32, 44]. In particular, to account for high rates of asymptomaticity among COVID-19 patients, three independent prior studies [16, 17, 20] proposed SEIR models with separate compartments for symptomatic and asymptomatic infections. We build on top of this work by proposing our own simplified SEIR model which is used by DOCTOR to design its optimal testing strategies.

Finally, we discuss related work on optimal intervention strategies, i.e., when/how to impose complete and partial lockdowns, etc., to contain COVID-19. There is some prior work on using Markovian models (e.g., MDPs, POMDPs) to design adaptive sequential strategies for imposing (and releasing) lockdown protocols depending on the current rate of COVID-19 spread [14, 36]. At the same time, Mikkulainen et al. [26] optimize intervention strategies using evolutionary optimization techniques. This body of research is complementary to our work as tackling COVID-19 effectively requires an optimal testing program applied in conjunction with an optimal intervention strategy.

## 3 THE OPTIMAL TESTING PROBLEM

We first describe our SEIR model and other preliminary notation that helps us define the optimal testing problem.

### SEIR Transmission Model

The SEIR model is a popular epidemiological model for simulating the progression of an epidemic through a population of individuals, and it has been used to successfully simulate the outbreak of many infectious diseases, e.g., Ebola [25], COVID-19 [23, 31], etc. In the standard SEIR model, a population of *N* individuals is split into four compartments: (i) Susceptible (**S**), i.e., individuals who have never been infected or exposed to the COVID-19 virus; (ii) Exposed (**E**), i.e., individuals who have been exposed to the virus, but are not infectious yet; (iii) Infectious (**I**), i.e., individuals who have been infected and can spread infection to other individuals; and (iv) Recovered (**R**), i.e., individuals who have recovered or died from the virus. Each of the four compartments represent a distinct phase in the progression of infectious diseases.

Further, to capture the most essential characteristics of COVID-19 transmission, we made two major adaptations to the standard SEIR model: (i) similar to He et al. [20], the **I** class is split into I_1_ and I_2_, which represents asymptomatic and symptomatic infected patients, respectively; (ii) we introduce a new compartment class named Hospitalization/Quarantine (**H/Q**), which represents individuals who are either hospitalized or are observing strict quarantine orders. Our two adaptations are necessary for modeling COVID-19 as (i) there is a high asymptomatic rate of COVID-19 infections [6, 7, 29], which can not be distinguished by the single **I** class in the standard SEIR model; and (ii) introducing the **H/Q** compartment enables us to model infected individuals who do not spread infection to anybody else (either because they are hospitalized or they observe strict stay-at-home quarantine orders).

Our SEIR model dynamics proceed in a series of discrete time steps. Let there be a population of *N* individuals that undergo SEIR model transmission dynamics. Let *T* denote the number of time steps for which the SEIR model dynamics are allowed to run. Each individual *n ϵ* {1, *N*} belongs to exactly one compartment at time *t* = 0. At each time *t ϵ* {1, *T*}, individuals in each compartment ‘flow’ to the next adjacent compartment at pre-determined rates. The flow dynamics of our model are explained in Figure 1. In particular, individuals in **S** move to **E** at a rate *α*_*s*_ →_*E*_, individuals in **E** move to I_1_ and I_2_ at rates 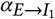 and 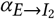, respectively. Similarly, individuals in I_1_ and I_2_ move to **R** at rates 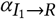 and 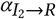. Also, individuals in **H/Q** move to **R** at a rate *α*_*H*/*Q* →*R*_. Finally, individuals in **R** do not take further part in transmission dynamics (i.e., we assume infected individuals upon recovery cannot be re-infected).

**Figure 1:**
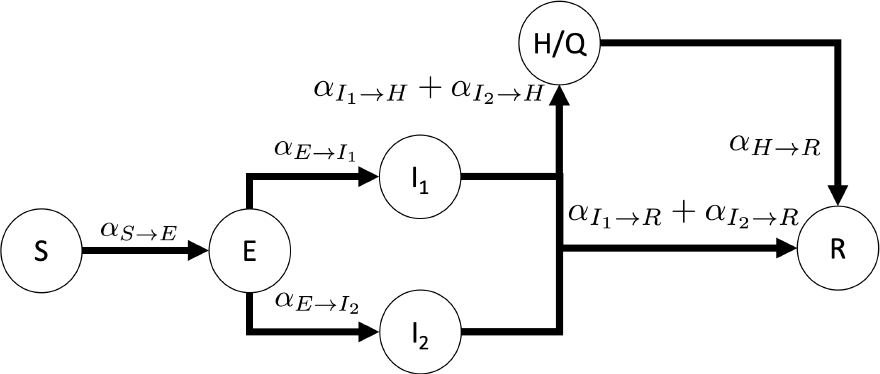
Flow Dynamics of our SEIR Model.

Our goal in the optimal testing problem is to minimize the cumulative number of individuals that are infected by COVID-19 (i.e., individuals in **I**_1_ and **I**_2_) across *T* time-steps of transmission. In order to achieve this goal, we need to formulate a sequential testing policy (formally defined below) that optimally allocates available testing kits to test symptomatic and asymptomatic individuals. We now elaborate on this distinction between conducting tests for symptomatic and asymptomatic individuals.

### Symptomatic VS Asymptomatic Testing

As part of the optimal testing policy, we assume that the policy maker is allowed to use his/her available COVID-19 testing kits to conduct two different kinds of tests: (i) Targeted Symptomatic Testing; and (ii) Random Asymptomatic Testing.

Targeted Symptomatic Testing (symptomatic testing, in short) focuses only on the people who exhibit COVID-19 symptoms and seek care at hospitals. The testing kits that are allocated for symptomatic testing would be distributed (in advance) across different hospitals. Only patients that show up at hospitals with COVID-19 like symptoms will be tested using these symptomatic testing kits. In our SEIR model, individuals in I_2_ can get tested using symptomatic testing kits (as all individuals in I_2_ are symptom-showing COVID-19 patients). In addition, we assume that a fraction of individuals in other compartments can suffer from Influenza Like Illnesses (ILI), and hence these ILI patients also show up at hospitals with COVID-19 like symptoms (we set this ILI fraction to 0.24 in our experiments, based on prior results [34]). Thus, in our SEIR model, these ILI patients can also be tested with symptomatic testing kits.

Unfortunately, symptomatic testing is not sufficient by itself, as (i) a large proportion of COVID-19 patients are asymptomatic (i.e., do not exhibit any symptoms), and hence they may never go to hospitals to get treated [6, 7, 29]. However, such asymptomatic patients can still spread the virus very rapidly among other individuals [3, 46]. (ii) Further, due to this large population of asymptomatic virus carriers, it is very difficult for epidemiologists and policy makers to understand where they are on the epidemic curve solely on the basis of symptomatic testing. In order to address this issue, we also consider Random Asymptomatic Testing as an option available to the policy maker. In our SEIR model, Random Asymptomatic Testing (asymptomatic testing, in short) focuses on all the individuals in **S, E**, I_1_, I_2_ and R, and randomly samples *m* individuals uniformly from these compartments to conduct COVID-19 tests on them (if the testing policy allocates *m* testing kits for asymptomatic testing).

Note that the positive COVID-19 diagnosis rate per test is much higher for symptomatic tests as compared to asymptomatic tests. Thus, while asymptomatic tests are essential to tackle infectious individuals in I_1_, they are not as efficient as symptomatic tests in discovering COVID-19 patients. In order to increase the efficiency of asymptomatic tests, we also incorporate group testing for asymptomatic tests in our optimal testing problem [38, 45].

We now formally define the optimal sequential testing policy. Our desired testing policy is adaptive, and we assume that we are allowed to make changes to our testing policy at *D* decision points (i.e., we are allowed to take *D* sequential actions). For example, *D* = 1 corresponds to a static testing policy, whereas if *D* = *T*, then we are allowed to change the testing policy at each time-step of the SEIR model. In practice, *D* is provided as input to the problem by policy makers who choose this value based on how fine-grained a policy they desire. Let *G*_0_ denote the policy maker’s initial understanding about the proportion of individuals in **S, E, I**_1_ and **R** (note that the policy maker can only perfectly observe the number of individuals in I_2_ and **H** /**Q**, but is uncertain about the number of individuals in the other compartments). Let ℬ {⟨*b*_1_, *b*_2_⟩ s.t. *b*_1_ ≥0, *b*_2_ ≥ 0. *b*_1_+ *b*_2=_ *U*_ca*p*_} denote the set of all possible ways in which *U*_*cap*_ testing kits can be divided among symptomatic and asymptomatic individuals, which represents the set of possible actions (choices) that can be taken by a policy maker at every decision point *d ϵ* {1,*D*}. Let *B*_*d*_, *ϵ* ℬ, ∀*d ϵ*{1, *D*} denote the policy maker’s action in the *d*^*th*^ time step. Upon taking action *B*_*d*_, the policy maker ‘*observes*’ the result of all COVID-19 tests that were conducted among symptomatic and asymptomatic individuals (as part of *B*_*d*_), and this updates their understanding of how many individuals remain in **S, E, I**_1_ and R. Let *G*_*d*_, ∀ *d ϵ*{1, *D*}denote the policy maker’s understanding resulting from *G*_*d*_ −_1_ with *observed* COVID-19 test result information from tests conducted in *B*_*d*_. Formally, we define a history *H*_*d*_, ∀ *d ϵ* {1, *D*}of length *d* as a tuple of past choices and observations *H*_*d*_ = ⟨*G*_0_, *B*_1_, *G*_1_, *B*_2_, .., *B*_*d*_ −_1_, *G*_*d*_ ⟩. Denote by ℋ_*d*_ = {*H*_*k*_ s.t. *k* ≤*d*}the set of all possible histories of length less than or equal to *d*. Finally, we define a *d*-step policy П_*d*_ : ℋ _*d*_ → ℬas a function that takes in histories of length less than or equal to *d* and outputs an action *B ϵ* ℬfor the current time step. We now formally define the optimal testing problem.

#### Problem 1. Optimal Testing Problem

*Given as input G*_0_, *integers N, T, D, and U*_*cap*_, *and SEIR model flow parameter values (as defined above). Denote by* ℛ (*H*_*D*_, *B*_*D*_) *the expected cumulative number of individuals in* **I**_1_ *and* **I**_2_ *at the end of D decision points, given the D-length history of previous observations and actions H*_*D*_, *along with B*_*D*_ *(i*.*e*., *the action chosen at decision point D). Let* 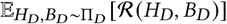 *denote the expectation over the random variables H*_*D*_ = ⟨*G*_0_, *B*_1_, .., *B*_*D*−1_, *G*_*D*_ ⟩ *and B*_*D*_, *where B*_*d*_ *are chosen according to* П_*D*_ (*H*_*d*_), ∀ *d ϵ* {1, *D*}, *and G*_*d*_ *are drawn according to distribution described by G*_*d*_ −_1_ *(i*.*e*., *the observation that we get when we execute B*_*d*_ *). The objective of the optimal testing problem is to find an optimal D-step policy* 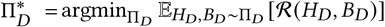.

## 4 DOCTOR POMDP

We cast the optimal testing problem as a POMDP because of three reasons. First, we have partial observability of the sizes of the S, E, I_1_ and R compartments in the optimal testing problem (similar to POMDPs). Second, similar to sequential POMDP actions, we are allowed to make *D* sequential changes to the testing policy (one change per each of the *D* decision points). Finally, POMDP solvers have recently shown great promise in generating near-optimal policies efficiently [43]. We now explain how we map the optimal testing problem into a POMDP.

### States

A POMDP state in our problem is a tuple *s* = ⟨**S, E**, I_1_, **I**_2_, **H Q, R**⟩, where variables S, E, I_1_, I_2_, H Q and R denote the number of individuals present inside the corresponding compartments of the SEIR model. For a POMDP state to be valid, we require **S + E + I**_**1**_ **+ I**_**2**_ **+ H + R** = *N*. Our POMDP has 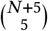states.

### Actions

At each decision point *d ϵ* {1,D} a total of, the policy maker has a total of*U*_*cap*_ COVID-19 testing kits that need to be allocated for testing individuals. An action inside our POMDP is a tuple *a* = ⟨*b*_1_, *b*_2_⟩, s.t. *b*_1_ ≥0, *b*_2_ ≥0 and *b*_1_+ *b*_2_ = *U*_*cap*_. Intuitively, *b*_1_ and *b*_2_ represent the number of testing kits that have been allocated for testing symptomatic and asymptomatic individuals, respectively. Our POMDP has *U*_*cap*_ + 1 different actions.

### Observations

Upon taking a POMDP action, we assume that the policy maker can “observe” the COVID-19 test results (positive or negative) of all individuals who were tested as part of the POMDP action. Formally, upon taking action *B*_*d*_ at decision point *d*, the POMDP observation is denoted as a binary vector (of length (*U*_*cap*_) 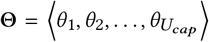. The variable *θ*_*i*_ = 1, ∀*i ϵ* {1, *U*_*cap*_} represents whether the *i*^*th*^ individual (who was tested in POMDP action *B*_*d*_) was diagnosed with COVID-19 (*θ*_*i*_ = 1) or not (*θ*_*i*_ = 0). Our POMDP has 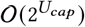 observations.

### Rewards

The cost *R* (*s, a, s*’) of taking action *a* in state *s* to reach state *s*’ is the number of active infected individuals in state *s*’. Over *D* decision points, DOCTOR’s cost function serves as a proxy for minimizing the cumulative number of COVID-19 infections.

### Transition & Observation Probabilities

Computation of exact transition and observation probability matrices (*T (s*^′^|*s, a*) and *O (o|a, s*^′^), respectively) is infeasible in our POMDP because these matrices are prohibitively large (due to large sized state, action and observation spaces). Therefore, we follow the paradigm of largescale online POMDP solvers [13, 33] by using a generative model Λ(*s, a*) ∼ (*s*’, *o, r*) of the transition and observation probabilities. This generative model allows us to generate on-the-fly samples from the exact distributions *T (s*’ |*s, a* and Ω (*o*| *a, s* ^′)^at low computational costs. Given an initial state *s* and an action *a*, our generative model Λ simulates the random process of SEIR model dynamics (as explained in Figure 1) to generate a random new state *s*’, an observation *o* and the obtained reward *r*. Simulation is done by “*playing*” out our SEIR model to generate sample *s*’. The observation sample *o* is then determined from *s*’ and *a*. Finally, the reward sample *r* depends on the number of active infected COVID-19 patients in s’ (as defined above). This simple design of the generative model allows significant scale and speed up.

### Initial Belief State

In our experiments, we initialize the belief state to be as close as possible to the real-world. In particular, the initial belief state is uniformly distributed over all POMDP states *s* in which **I** is set to the current number of COVID-19 infections in the population of interest. Then **I**_1_ and **I**_2_ are split from I based on COVID-19 asymptomatic rate ϕ (we experiment with different ϕ values). For example, if we instantiate a SEIR model for Santiago, the initial belief state contains all states in which I is equal to current number of active cases in Santiago. And if we assume the ϕ = 0.7, then |**I**_1_ | = 0.7 I, and |**I**_2_ | = 0.3 |**I** |.

In this paper, we solve the DOCTOR POMDP model using POMCP, a well-known online POMDP solver that relies on Monte Carlo tree search to find near-optimal online policies. For completeness, we provide a brief overview of the POMCP algorithm.

### POMCP

POMCP [33] uses UCT based Monte-Carlo tree search (MCTS) [5] to solve POMDPs. At every stage, given the current belief state *ϵ*, POMCP incrementally builds a UCT tree that contains statistics that serve as empirical estimators (via MC samples) for the POMDP Q-value function 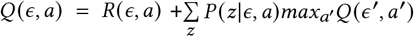. The algorithm avoids expensive belief updates by maintaining the belief at each UCT tree node as an unweighted particle filter (i.e., a collection of all states that were reached at that UCT tree node via MC samples). In each MC simulation, POMCP samples a start state from the belief at the root node of the UCT tree, and then samples a trajectory that first traverses the partially built UCT tree, adds a node to this tree if the end of the tree is reached before the desired horizon, and then performs a random rollout to get one MC sample estimate of *Q ϵ, a*. Finally, this MC sample estimate of *Q (ϵ, a*) is propagated up the UCT tree to update Q-value statistics at nodes that were visited during this trajectory. Note that the UCT tree grows exponentially large with increasing state and action spaces. Thus, the search is directed to more promising areas of the search space by selecting actions at each tree node *h* according to the UCB1 rule [24], which is given by: 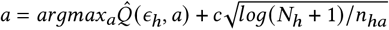. Here, 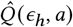 resents the Q-value statistic (estimate) that is maintained at node *h* in the UCT tree. Also, *N*_*h*_ is the number of times node *h* is visited, and *n*_*ha*_ is the number of times action *a* has been chosen at tree node *h* (POMCP maintains statistics for *N*_*h*_ and *n*_*ha*_ ∀ *a* ϵ *A* at each tree node *h*). We use POMCP to solve DOCTOR’s POMDP model.

## 5 EXPERIMENTAL RESULTS

We provide a comprehensive evaluation of the strengths and weaknesses of DOCTOR’s testing policy under a wide variety of conditions. We provide four sets of simulation results. First, we evaluate DOCTOR’s effectiveness in controlling the spread of COVID-19 by applying its testing strategy (in simulation) to the city of Santiago, Panama (a country with the world’s highest COVID-19 infections per capita [1]). Second, we analyze whether the performance gains achieved by DOCTOR’s testing strategy are robust to variations across a multitude of parameter values. Third, we carefully analyze key characteristics of DOCTOR’s testing policy to gain unique insights about what DOCTOR has learnt about this problem domain (of finding optimal testing strategies), and to understand whether these insights can be translated into actionable lessons for policy makers as they design testing strategies for COVID-19. Finally, we characterize conditions (of the COVID-19 epidemic) under which it would be most beneficial for governments to use DOCTOR’s testing strategies. Specifially, we provide an *easy-hard-easy* computational pattern characterization of optimal testing problems, and show that DOCTOR’s performance gains are largest for hard problems, whereas the gains diminish on comparatively easier problems.

All our experiments are run on a 2.8 GHz Intel Xeon processor with 256 GB RAM. All experiments are averaged over 100 runs. In all experiments, we use a default value of *N* = 89, 000 (which is Santiago’s population [40]), *D* = 30 and *T* = 30 (unless specified otherwise). Based on findings in [38, 45], COVID-19 test sensitivity and specificity values are set to 0.90 and 0.99, respectively. Also, we set a default budget constraint of *U*_*cap*_ = 500 testing kits per decision point in our experiments (unless specified otherwise). The POMCP-based DOCTOR model is run with 2^10^ Monte-Carlo simulations at each decision point. Further, in all our experiments, we use results from [15] to instantiate our SEIR model with the following values of flow rates: 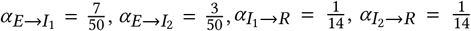, and 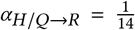. In particular, the value of *α*_*s*_ →_*E*_ is dependent on the basic reproduction number of COVID-19 (*R*_0_) and the number of individuals in I, which is denoted as 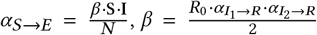, where *β* represents the COVID-19 transmission rate. All experiments are statistically significant under bootstrap-t (*p* = 0.05).

### Baselines

We compare DOCTOR against four different baseline testing strategies. We use (i) 100% symptomatic testing (SY in the figures), i.e., allocate all available testing kits to symptomatic individuals at each decision point. We use SY as a baseline as this has been the primary testing strategy used by Panama’s government until now, e.g., Panama had not tested asymptomatic individuals until 4^*th*^ September, 2020 [30]. Using this baseline allows us to compare DOCTOR with a real-world government’s effort (in simulation). Next, we use (ii) 100% asymptomatic testing (ASY), i.e., allocate all available testing kits to asymptomatic individuals; (iii) 50% symptomatic and 50% asymptomatic testing (50-ASY), i.e., equally divide testing kits among symptomatic and asymptomatic individuals; and finally (iv) a uniform random testing policy (Random), i.e., select a random testing action *B*_*d*_ at every decision point *d ϵ D*.

### 5.1 DOCTOR’s performance in Panama

First, we evaluate the performance of DOCTOR’s testing policy against all other baselines, when applied to the city of Santiago, the 5^*th*^ largest city in Panama. Since city-level COVID-19 case information is not available for Panamanian cities, we initialize Santiago’s SEIR model using Panama’s country-level COVID-19 case information. In particular, we set the initial SEIR compartment proportions to ⟨S = 97.47%, E = 0.27%, I_1_ = 0.45%, I_2_ = 0.19%, R = 1.62%, H/ Q = 0%⟩, which matches the COVID-19 infection numbers in Panama on 2^*nd*^ September, 2020. Note that **H** /**Q** is set to zero because we only count **H** /**Q** from the beginning of the testing period. Next, DOCTOR and the other baselines were used to solve an optimal testing problem (defined according to this instantiated SEIR model and the other parameter values described above).

Figure 2 compares the result of *executing* DOCTOR’s testing policy against baselines by tracking the evolution of the underlying SEIR model over *D* = 30 decision points. Figures 2(a), 2(b), 2(c) and 2(d) show the progression in the sizes of S, I, I_1_ and I_2_ compartments of the SEIR model (respectively) over *D* = 30 decision points. The X-axis in these figures represents the different decision points, and the Y-axis shows the size of the different compartments. For example, DOCTOR’s testing strategy achieved a size of| S |= 85, 528, |I |= 136, |I_1_| = 120 and |I_2_| = 16 after the 30^*th*^ decision point.

**Figure 2:**
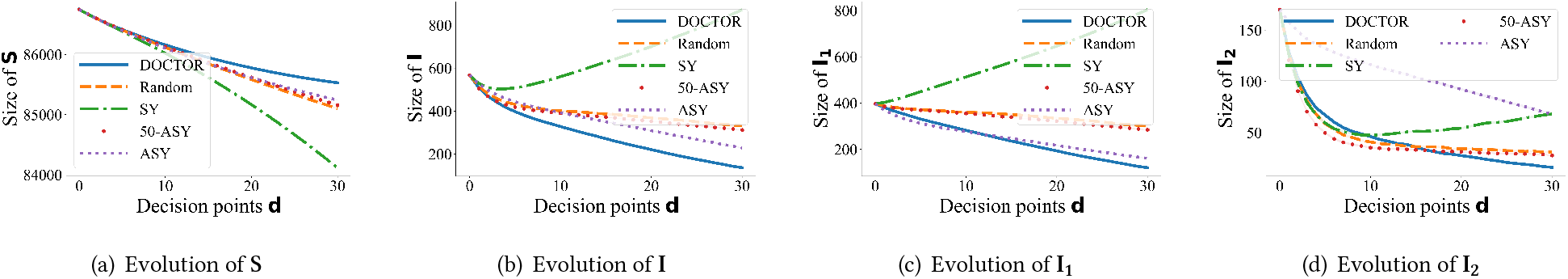
Evaluating DOCTOR’s performance in Panama.

Figure 2(b) shows that DOCTOR significantly outperforms all baselines - its testing strategies result in ∼ 40% fewer COVID-19 infections by the 30^*th*^ decision point (as compared to ASY, the next best performing baseline). Further, this figure shows that SY performs very poorly - it performs ∼ 60% worse than Random and ASY-50, and it leads to a ∼ 550% increase in COVID-19 infections over DOCTOR. This establishes the superior performance of DOCTOR over SY (and other baselines), which illustrates the potential benefits of using DOCTOR’s adaptive strategy in Panama.

Figures 2(c) and 2(d) provide a preliminary insight into how DOCTOR achieves significant reductions in the number of COVID-19 infections. Specifically, these figures show why baseline testing strategies fail: (i) ASY performs only ∼ 30% worse than DOCTOR in minimizing asymptomatic infections |I_1_ |, but performs ∼ 300% worse than DOCTOR in minimizing symptomatic infections |I_2_ | (since ASY only tests asymptomatic individuals). (ii) On the other hand, SY performs ~ 300% worse than DOCTOR in minimizing |I_2_|, and performs ~ 550% worse than DOCTOR in minimizing I_1_ (since SY only focuses on symptomatic individuals). (iii) ASY-50’s behavior is not as extreme as SY (in Figure 2(c)) or ASY (in Figure 2(d)), yet it performs worse than DOCTOR due to its lack of adaptivity. (iv) DOCTOR is the only strategy which intelligently minimizes both |I_1_| and |I_2_| by adaptively changing the allocation of testing kits according to the stage of the epidemic. (v) Further, DOCTOR’s testing strategy results in the largest S at the end of the 30^*th*^ decision point (Figure 2(a)), which illustrates DOCTOR’s (relative) success in preventing susceptible individuals in S from getting infected. These figures show that at least in simulation, DOCTOR was highly effective in controlling the number of COVID-19 infections in Santiago.

### 5.2 Is DOCTOR robust to varying parameters?

Having illustrated DOCTOR’s effectiveness in controlling COVID-19 infections in Santiago, we now analyze if DOCTOR’s superiority over baselines is robust to different initial parameter values.

Figures 3(a), 3(b), 3(c) and 3(d) compares the percentage improvement achieved (in terms of |I| = |I_1_ | + |I_2_ |) by DOCTOR over other baselines with varying values of *N*, COVID-19 asymptomatic rate (ϕ), test sensitivity (*δ*), and pooling sizes for group testing (*ρ*), respectively. The Y-axes in these figures show the percentage improvement achieved by DOCTOR over other baselines, whereas the X-axes show the varying parameter values. For example, when *N* = 100, 000 (in Figure 3(a)), DOCTOR outperforms ASY, ASY-50, SY and Random by 55%, 83%, 53% and 36%, respectively. Figure 3(a) shows that (i) DOCTOR consistently outperforms all other baselines; and (ii) DOCTOR’s superior performance does not diminish at higher/lower values of *N*, which illustrates that DOCTOR is robust to variation in *N*.

**Figure 3:**
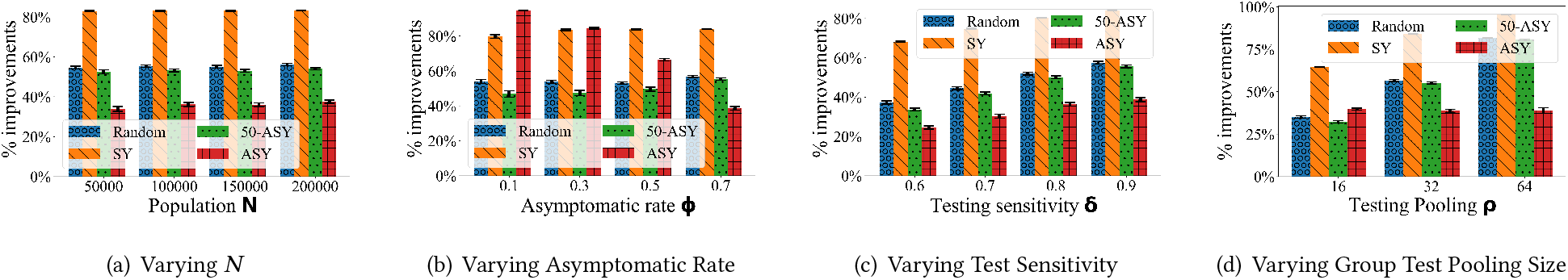
Minimal impact on DOCTOR’s performance (over baselines) with variation in parameter values.

Next, in order to account for the large variance in COVID-19 asymptomatic rate (ϕ), test sensitivity (*δ*) and pooling size (*ρ*) values reported in prior literature [6, 7, 29, 38, 45, 47], we experimentally verify that DOCTOR’s superior performance over baselines does not lie on a knife’s edge in terms of the ϕ, *δ* and *ρ* values that are used inside DOCTOR. Figure 3(b) shows that (i) ASY’s performance improves significantly with increasing values of *ϕ*. This is reasonable because when a large fraction of infected individuals are asymptomatic, it is (near) optimal to allocate all testing kits for them. (ii) Even at large *ϕ* values, DOCTOR outperforms ASY by 38%. (iii) DOCTOR consistently outperforms all other baselines, and its performance does not diminish against these other baselines at larger/smaller *ϕ*. Thus, Figure 3(b) shows that DOCTOR is mostly robust to variation in *ϕ* (except against ASY, which becomes near optimal at high *ϕ* values).

Figure 3(c) shows that (i) DOCTOR’s performance improvement (over baselines) increases with increasing values of *δ*. This is reasonable as high *δ* values mean that DOCTOR has less uncertainty to contend with, which leads to better high-quality solutions. (ii) Even at low test sensitivity values (e.g., *δ* = 0.6, as reported in [47]), DOCTOR outperforms ASY (the next best baseline) by 24%. This shows that DOCTOR is robust to variation in *δ*.

Finally, Figure 3(d) shows that (i) varying *ρ* minimally impacts DOCTOR’s performance improvement over ASY (the best performing baseline); and (ii) all other baselines exhibit degraded performance at higher values of *ρ*. In summary, Figure 3 shows that across a wide range of parameter values, DOCTOR exhibits 30% average performance improvement against other baselines, which illustrates its robustness to changes in these parameter values.

### 5.3 What did DOCTOR learn?

Having illustrated DOCTOR’s robustness to varying initial conditions, we now analyze its outputted testing policy to gain insights about the reasons behind the optimality of DOCTOR’s policy.

Figures 6(a) and 6(b) illustrate the testing policy output by DOCTOR over 30 decision points with *U*_*cap*_ = 300 and *U*_*cap*_ = 500, respectively. The X-axis shows the different decision points, whereas the Y-axis illustrates the action chosen by DOCTOR. For example, in Figure 6(a), DOCTOR assigned 1 ∼ 95 (and ∼ 105) tests to symptomatic (and asymptomatic) individuals at the 1^*st*^ decision point. These figures reveal a crucial insight about DOCTOR’s policies.

Specifically, DOCTOR’s outputted testing policies proceed in two sequential phases. In the first phase (during the initial few decision points), DOCTOR spends more effort in testing symptomatic individuals, e.g., DOCTOR allocates ∼65% of its available testing kits for symptomatic individuals at the 1^*st*^ decision point in Figure 6(a). Over time, as the number of symptomatic individuals diminishes (due to DOCTOR’s emphasis on testing such individuals), DOCTOR begins its second phase of testing. In this second phase, DOCTOR switches its attention towards asymptomatic testing, and gradually increases the number of testing kits allocated to asymptomatic testing as decision points proceed. For example, the proportion of tests allocated to asymptomatic testing increases from ~35% at the 1^*st*^ decision point to ∼80% at the 30^*th*^ decision point (Figure 6(a)).

This sequential two-phased nature of testing is a defining characteristic of testing policies output by DOCTOR, and is a key differentiating factor between DOCTOR’s optimal strategies and other baselines. In addition, DOCTOR needs to account for several other considerations in finding its optimal policy: (i) what proportion of resources to invest in symptomatic testing in the 1^*st*^ phase, (ii) at what decision point should the 2^*nd*^ phase begin, (iii) at what rate should the number of resources allocated to asymptomatic testing be increased in the 2^*nd*^ phase. DOCTOR answers all these questions via POMDP style look ahead search.

This insight about DOCTOR’s optimal testing strategy has profound policy implications for COVID-19 testing in the real-world. Even though it might be infeasible for governments of developing countries to formulate their COVID-19 testing policy by running computationally heavy POMDP programs, our insights about the superior performance of two-phased testing policies suggest that governments should consider investing in testing policies which (i) start by focusing on symptomatic testing (within the means of available testing capacity), and (ii) when symptomatic cases start falling, the policy should switch over gradually to focus more on conducting random asymptomatic testing.

Next, we also analyze the impact of varying parameter values on the testing policies output by DOCTOR. To understand the next set of experiments, we formally define the “average” POMDP action chosen by DOCTOR’s testing policy over 30 decision points. With *U*_*cap*_ testing kits available per decision point, let the actions chosen by DOCTOR over 30 decision points be 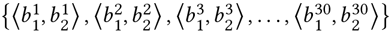 . Let 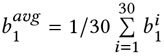 be the average number of symptomatic tests recommended by DOCTOR’s policy (per decision point). Then, the “average” POMDP action (which we represent in Figure 4) is 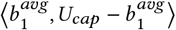.

**Figure 4:**
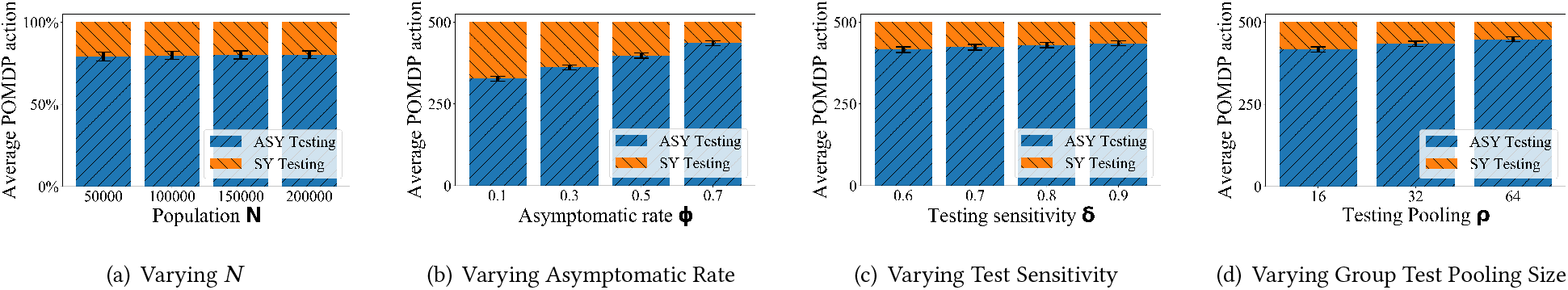
Impact of changing parameter values on DOCTOR’s testing strategy.

Figures 4(a), 4(b), 4(c) and 4(d) illustrate DOCTOR’s testing policy with varying values of *N, ϕ, δ* and *ρ*, respectively. The Y-axes in these figures show the “average” POMDP action chosen by DOCTOR, and the X-axes show varying parameter values.

Figures 4(a) and 4(c) show that ceteris paribus, DOCTOR’s testing policy is minimally affected (in average terms) by changing values of *N* and *δ*. Further, Figure 4(b) shows that upon increasing *ϕ*, DOCTOR allocates an increasing fraction of available testing kits for asymptomatic individuals, e.g., the proportion of tests for asymptomatic individuals increased from 65% (*ϕ* = 0.1) to 87% (*ϕ* = 0.7). This is expected behavior because with increasing rates of asymptomaticity, the number of asymptomatic individuals increases, and as a result, random testing of asymptomatic individuals leads to greater utilities. Similarly, Figure 4(d) shows that with increasing values of *ρ*, DOCTOR marginally increases the fraction of testing kits allocated for asymptomatic testing. Again, this is expected behavior because increasing pooling sizes increases the per-test efficiency of conducting tests on asymptomatic individuals, and as a result, DOCTOR allocates more tests for such individuals.

### 5.4 When to use DOCTOR?

In this last part of our evaluation, we characterize the conditions in which it is advisable for resource-strapped governments to rely on DOCTOR to determine their COVID-19 testing policy. At a high level, we observe that DOCTOR’s suitability for use often depends on the stage of the (COVID-19) epidemic at which the new testing policy is implemented. While we find that DOCTOR’s testing policy achieves the best performance irrespective of the stage at which it is implemented, we surprisingly find that DOCTOR’s superiority over baselines diminishes when its testing strategy is implemented with either extremely high or extremely low epidemic severity, which means DOCTOR’s superiority is most pronounced during the intermediate severity of the epidemic. We illustrate this interesting phenomenon below.

In order to model different stages of the epidemic, we vary the values of |I| /*N* in our initial SEIR compartment partition, i.e., we vary the fraction of individuals who belong to the I compartment before any testing strategy is employed. For simplicity of presentation, we only compare DOCTOR against the best performing baseline in Figure 5. Note that the best performing baseline can be different at different parameter values. Figure 5(a) compares the percentage improvement achieved by DOCTOR over the best performing baseline with varying values of |I |/*N* in the initial SEIR compartment partition. The X-axis shows increasing values of I *N*, and the *Y*-axis shows percentage improvement achieved by DOCTOR over the best performing baseline. This figure shows that DOCTOR outperforms the best performing baseline by 40% when |I|/ *N* = 0.005 (i.e., intermediate severity). Surprisingly, DOCTOR’s performance improvement diminishes at extreme values of |I|/ *N*, which represent different high and low epidemic severities.

**Figure 5:**
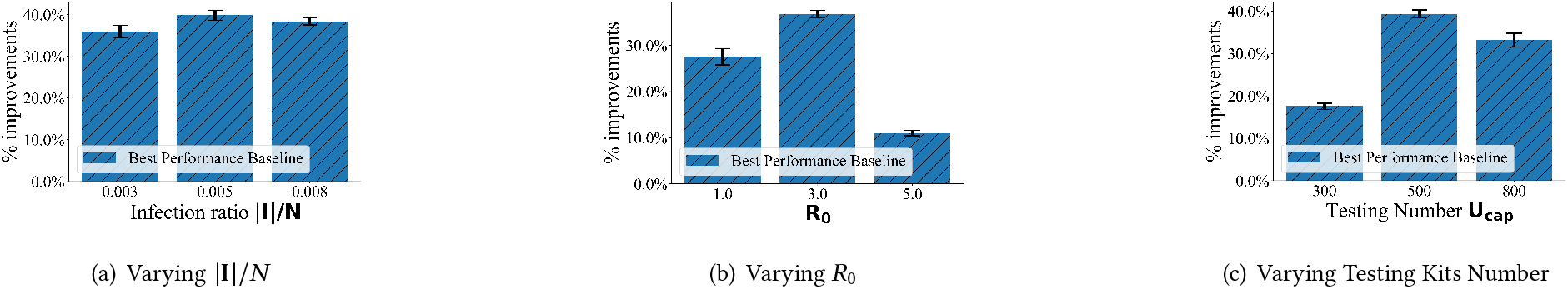
Reduction in performance gains achieved by DOCTOR (over baselines) at extreme parameter values.

**Figure 6:**
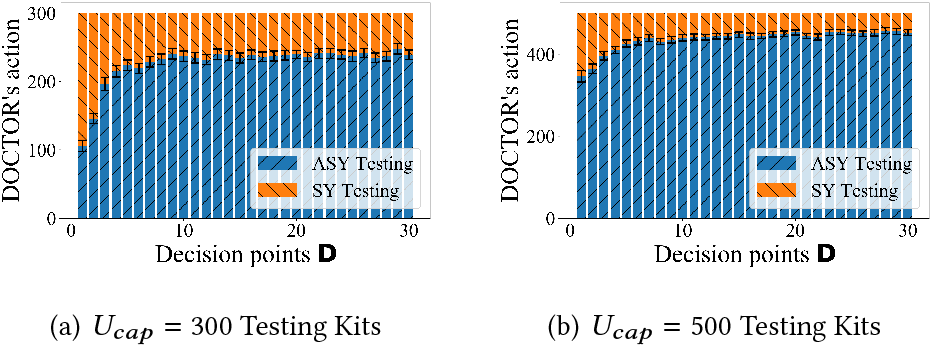
Visual representation of DOCTOR’s testing strategy over 30 decision points.

Upon closer examination, this makes sense as solving the optimal testing problem at extremely low (i.e., small |I| / *N*) or extremely high (i.e., large |I|/ *N*) epidemic severity represent “*easier*” problems to solve from a computational perspective. This is because at small values of |I|/ *N*, very few individuals are infected, as a result, almost any testing strategy can work well at this stage. Similarly, at large values of |I|/ *N*, the epidemic has already infected a large proportion of individuals, and beyond this point, almost every testing strategy (including DOCTOR) will lead to a poor performance. On the other hand, solving the optimal testing problem with intermediate |I| /*N*, represents “*harder*” computational problems because at this stage, the long-term trade off between investing resources in asymptomatic versus symptomatic testing is most apparent. As a result, we truly see the benefits of long-term look ahead search and policy optimization (techniques used by DOCTOR) when |I| /*N* is in the intermediate range.

Thus, we see an *easy-hard-easy* computational pattern to problems solved by DOCTOR with varying values of |I|/ *N*. In fact, we see this easy-hard-easy pattern with other parameters as well. Figure 5(b) compares the percentage improvement achieved by DOCTOR over the best performing baseline with varying values of *R*_0_. This figure shows that DOCTOR is only effective when *R*_0_ is neither too low nor too high. For example, when *R*_0_ = 2.0, DOCTOR achieves a ~40% smaller I compartment as compared to baselines. However, when *R*_0_ = 1.0 (or *R*_0_ = 5.0), DOCTOR’s improvement over baselines diminishes to 28% (or 11%). Again, this is reasonable because smaller and larger *R*_0_ values correspond to easier problems - the disease either does not spread fast (at small *R*_0_) or spreads too fast (at large *R*_0_), hence almost any testing strategy for reducing the infection number will perform well (for small *R*_0_) or perform bad (for large *R*_0_). It is only at intermediate *R*_0_ values (corresponding to hard problems) that the benefits of sequential adaptive optimization (i.e., DOCTOR) are realized.

Finally, we also observe this easy-hard-easy pattern when we vary the testing capacity *U*_*cap*_. Figure 5(c) compares the percentage improvement achieved by DOCTOR over the best performing baseline with varying values of *U*_*cap*_. This figure shows that DOCTOR is only effective when *U*_*cap*_ is neither too low nor too high. For example, when *U*_*cap*_ = 500, DOCTOR outperforms baselines by 39%. However, when *U*_*cap*_ = 300 (or *U*_*cap*_ = 800), DOCTOR’s improvement over baselines diminishes to 17% (or 28%). Intuitively, this makes sense as when you have a low capacity of testing (easy problem), it is optimal to allocate all those tests for symptomatic patients. In this case, DOCTOR is unable to outperform the SY baseline. Similarly, with very high testing capacities (easy problem), it is optimal to allocate all tests for asymptomatic testing. In this case, DOCTOR is unable to outperform the ASY baseline. It is only at intermediate *U*_*cap*_ values (hard problem) that DOCTOR is able to outperform all baselines significantly. In summary, Figures 5(a), 5(b) and 5(c) illustrate an interesting easy-hard-easy computational pattern to problems solved by DOCTOR, and show that DOCTOR’s performance gains are most pronounced on hard problems. Accordingly, this shows that it is advisable to use DOCTOR for determining testing strategies in the real-world only when |I | /*N, R*_0_ and *U*_*cap*_ values are in the intermediate range, which represent the intermediate severity of an epidemic. With extremely high or extremely low severity, simpler static baselines may be more preferable.

## 6 CONCLUSION

By evaluating DOCTOR in different real-world scenarios, we reveal the two-phased nature of DOCTOR’s testing policies. Additionally, we illustrate that the benefits of these policies are robust against variations across different input parameters. Surprisingly, we also discover that the benefits of DOCTOR’s policy show an *easy-hard-easy* computational pattern with some varying input parameters. Such easy-hard-easy patterns have also been observed in other problems, e.g., Security Games [22], 3-SAT [9, 27] and Influence Maximization [19]. In summary, DOCTOR represents a highly useful decision-aid for governments as they plan their COVID-19 testing strategies, especially during the intermediate stages of the epidemic. Specifically, DOCTOR’s testing strategies result in ∼40% fewer COVID-19 infections (thereby leading to lesser loss of life).

## Data Availability

Panama's COVID-19 infections number

https://www.worldometers.info/coronavirus/country/panama/

## Notes

### Competing Interest Statement

The authors have declared no competing interest.

### Clinical Trial

This paper is not related to clinical study.

### Funding Statement

No funding.

### Author Declarations

No approval needed.

